# HNF1B-MODY in the Norwegian MODY Registry and the Norwegian Childhood Diabetes Registry: Clinical insights and prevalence informed by genetic and functional evaluation

**DOI:** 10.64898/2026.03.23.26348894

**Authors:** Aishwarya Pavithram, Bente B. Johansson, Erling Tjora, Pernille Svalastoga, Khadra A. Mohamed, Ingvild L. Koløen, Maren Toftdahl, Torild Skrivarhaug, Marc Vaudel, Lise Bjørkhaug, Kristin A. Maloney, Toni I. Pollin, Stefan Johansson, Christine Bellanné-Chantelot, Jørn V. Sagen, Janne Molnes, Pål R. Njølstad

## Abstract

Interpreting *HNF1B* variants is challenging in clinical practice. We aimed to integrate functional, clinical, and family data to improve variant classification, describe clinical features of carriers and report registry-level prevalence of *HNF1B* alterations. Clinical, genetic, and family data were analyzed from the Norwegian MODY Registry (NMR) and the Norwegian Childhood Diabetes Registry (NCDR). Clinical features of sequence variant and 17q12 deletion (17q12del) carriers were summarized, and variants were classified using ACMG-AMP-ClinGen criteria. Registry-level prevalence was reported with 95% confidence intervals. *HNF1B* sequence variants were functionally assessed, showing that the lower transactivation (TA) was associated with higher clinical severity. Eleven variants demonstrated impaired functional activity, with TA inversely correlated with clinical burden (ϱ = −0.701, p = 0.002). We identified 28 individuals with 17q12del (21 in NMR, seven in NCDR) and 15 individuals carrying 14 unique (LP/P) sequence variants, all detected in the NMR. Overall, 36/486 probands (7.4%) with genetically confirmed monogenic diabetes in the NMR carried an LP/P *HNF1B* sequence variant or 17q12del. In the NCDR, ∼ 0.2% carried 17q12del (7/3,583; 3/7 GADA/IA-2A-positive). Functional data enabled reclassification of three variants. Since many pediatric 17q12del carriers in the NMR were referred for testing due to structural renal anomalies without diabetes, *HNF1B* screening should be considered in children with renal/extra-renal features, irrespective of diabetes or autoantibody status.

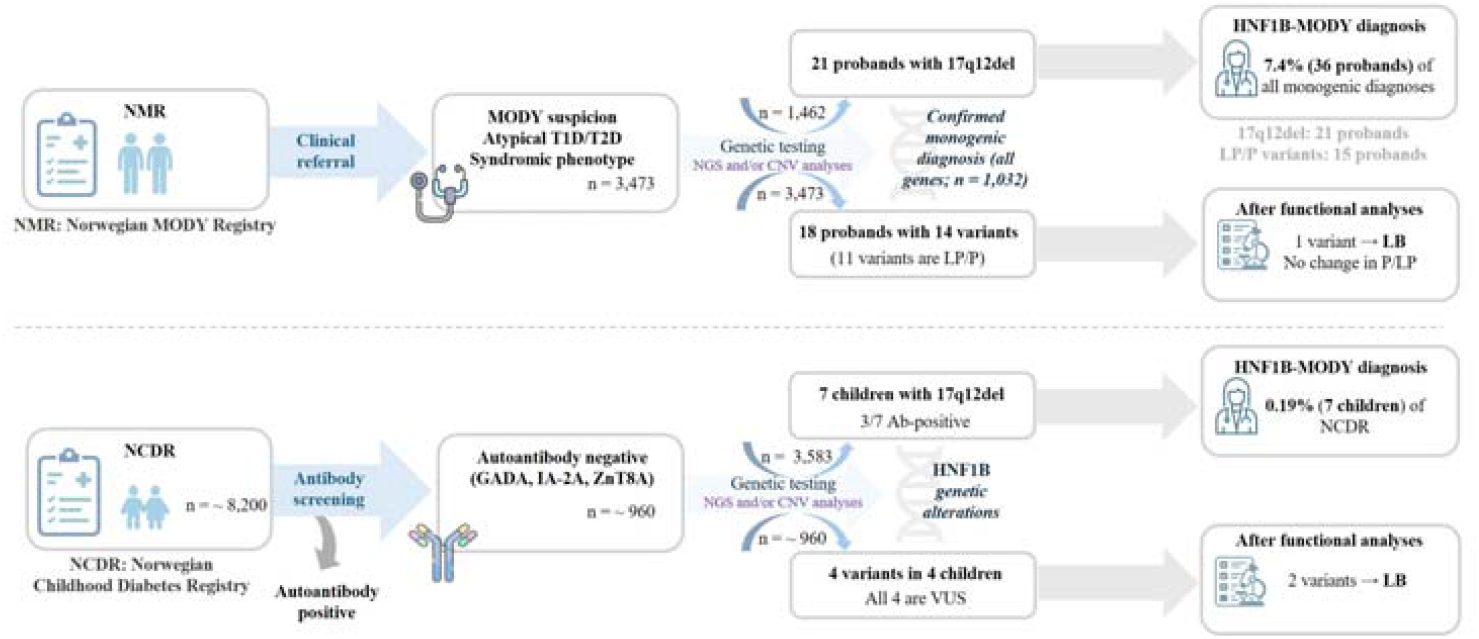

## 1. Introduction

Maturity-onset diabetes of the young (MODY) is a form of autosomal-dominant monogenic diabetes with early onset and clinical overlap with type 1 and type 2 diabetes[1-3]. MODY is caused by single-gene defects mostly affecting beta-cell development and function, with at least ten known causative genes[4]. Population studies have shown that pathogenic alterations in hepatocyte nuclear factor 1 alpha (*HNF1A*) and glucokinase (*GCK*) account for the largest proportion of MODY (∼ 30-50%), followed by *HNF4A* (∼ 10%) and *HNF1B* (∼ 5%)[5, 6]. In Norway, according to the Norwegian MODY registry (NMR), HNF1A-and GCK-MODY are most prevalent, followed by HNF1B-and HNF4A-MODY. Among established MODY subtypes, HNF1B-MODY is distinctive for its multisystemic presentation, combining diabetes with renal and genitourinary structural anomalies, hepatobiliary involvement, uric acid and electrolyte disorders, neurodevelopmental features, and pancreatic hypoplasia[7]. Mechanisms of disease include heterozygous loss of function (LoF) or functional impairments, either from 17q12 deletions (17q12del; haploinsufficiency) that encompass the *HNF1B* gene, or sequence variants affecting functional domains of the protein (dimerization domain (DD), DNA-binding domain (DBD), and transactivation domain (TAD)). Despite clear gene-disease association, the interpretation of *HNF1B* missense variants remains difficult. Phenotypic heterogeneity blurs genotype-phenotype correlations[8], the high rate of *de novo* occurrences decreases the ability to perform segregation analyses, and functional data for missense variants are limited. Moreover, incomplete penetrance and variable expressivity of associated manifestations complicate the interpretation[8-10]. Altogether, this may result in inconsistent variant interpretation and possible misdiagnosis.

Therefore, the present study focuses on clinical phenotyping of both sequence variants and 17q12del cases, with additional functional evaluation for sequence variants. Specifically, we investigated the NMR and the Norwegian Childhood Diabetes Registry (NCDR) to: (i) describe and compare clinical features among sequence variant and 17q12del carriers; and (ii) integrate functional readouts with clinical and family data to improve variant interpretation. By obtaining a precise variant classification, we can not only provide improved treatment and genetic counselling for individuals but also obtain a more accurate estimate of HNF1B-MODY prevalence. Thus, extending our NCDR-based *HNF1A* framework (lower-bound prevalence ∼ 0.34%)[3, 11-13] to *HNF1B*, we aim to precisely classify variants using functional analyses to reduce variants of uncertain significance (VUS) burden, and estimate registry-level prevalence of HNF1B-MODY in NMR and NCDR.

## 2. Results

### 2.1. Study cohort and *HNF1B* alterations

Among individuals referred for genetic testing in the NMR, 1,032 of 3,473 (30%) have received a confirmed genetic diagnosis of monogenic diabetes (Figure 1). In this study, across two registries we identified 17 unique *HNF1B* sequence variants among 18 participants (14 in the NMR and four in the NCDR) (Figure 2). One variant (p.S362F) was identified in individuals of both registries. The p.S362F carrier in the NMR cohort developed diabetes at the age range of 16-21 years, while the child in the NCDR cohort was diagnosed with diabetes at the age range of 1-5. First-degree relatives of both individuals were unaffected, and the variant is currently classified as VUS. All (likely) pathogenic (LP/P) variant carriers were identified in the NMR, whereas all variants identified in individuals of the NCDR were classified as likely benign (LB) or VUS. In addition, 28 individuals (21 probands in the NMR and seven children in the NCDR) were found to carry 17q12del. The majority of 17q12del carriers were identified in the NMR (81%), although one in five were identified through the NCDR (19%). No sex difference was found within and between groups (Table 1). Median age at referral was comparable between LP/P sequence variant carriers and 17q12del carriers (29.0 [IQR 15.5–44.0] vs 34.0 [IQR 11–41] years; Mann–Whitney U = 589.5, p = 0.513).

**Table 1.** Summarized clinical characteristics of all LP/P sequence variants and 17q12del carriers.

| Characteristics | Total | LP/P variant carriers | Total | 17q12del carriers |
| --- | --- | --- | --- | --- |
| N (all carriers) | — | 35 |  | 37 |
| Female (%) | 35 | 18 (51) | 37 | 20 (54) |
| NMR (%) | 35 | 35 (100) | 37 | 30 (81) |
| NCDR (%) | — | — | 37 | 7 (19) |
| Age at referral, years (IQR) | 35 | 29.0 (15.5-44.0) | 37 | 34 (11-41) |
| Diabetes-present (%) | 34 | 25 (74) | 37 | 25 (68) |
| DM/renal disease in first degree relatives (%) | 31 | 29 (94) | 22 | 22 (100) |
| Age at diabetes diagnosis, years (IQR) | 21 | 25 (16.0-36.0) | 20 | 27.5 (17.7-36.2) |
| BMI at referral, kg/m<sup>2</sup> (IQR) | 19 | 22.6 (18.8-24.3) | 20 | 22 (21.0-24.4) |
| HbA1c at referral, % (IQR) | 28 | 6.95 (5.8-8.15) | 24 | 7 (6.1-8.1) |
| HbA1c, mmol/mol (IQR) | 28 | 52.5 (40-65.5) | 24 | 52.5 (43.2-65.7) |
| Insulin, at referral (%) | 28 | 19 (68) | 27 | 13 (48) |
| **HNF1B-MODY features** |  |  |  |  |
| Renal disease (%) | 22 | 22 (100) | 18 | 18 (100) |
| Electrolyte or uric acid imbalance (%) | 3 | 3 (**100**) | 8 | 8 (100) |
| Pancreas structural abnormality or exocrine pancreatic insufficiency (%) | 14 | 11 (78) | 4 | 4 (**100**) |
| Genital tract abnormality (%) | 2 | 2 (**100**) | 2 | 2 (**100**) |
| Liver test abnormality, cholestasis (%) | 3 | 3(**100**) | 2 | 2 (**100**) |
| Gout and Hyperparathyroidism (%) | 1 | 1 (**100**) | — | — |
| Intellectual/Learning disability (%) | — | — | 4 | 4 (**100**) |

Data are presented as median (interquartile range [IQR], 25th–75th percentile) for continuous variables and as n (%) for categorical variables. For each characteristic, n indicates the number of individuals with available data (denominator shown in the “Total” column). Percentages were calculated using the number with available data as the denominator. Both probands and affected family members carrying LP/P sequence variants or 17q12del were included. Items marked with \*\* indicate very small denominators ( $n \leq 4$ ). LP/P = likely pathogenic/pathogenic; del = deletion; NMR = Norwegian MODY Registry; NCDR = Norwegian Childhood Diabetes Registry

**Figure 1.**
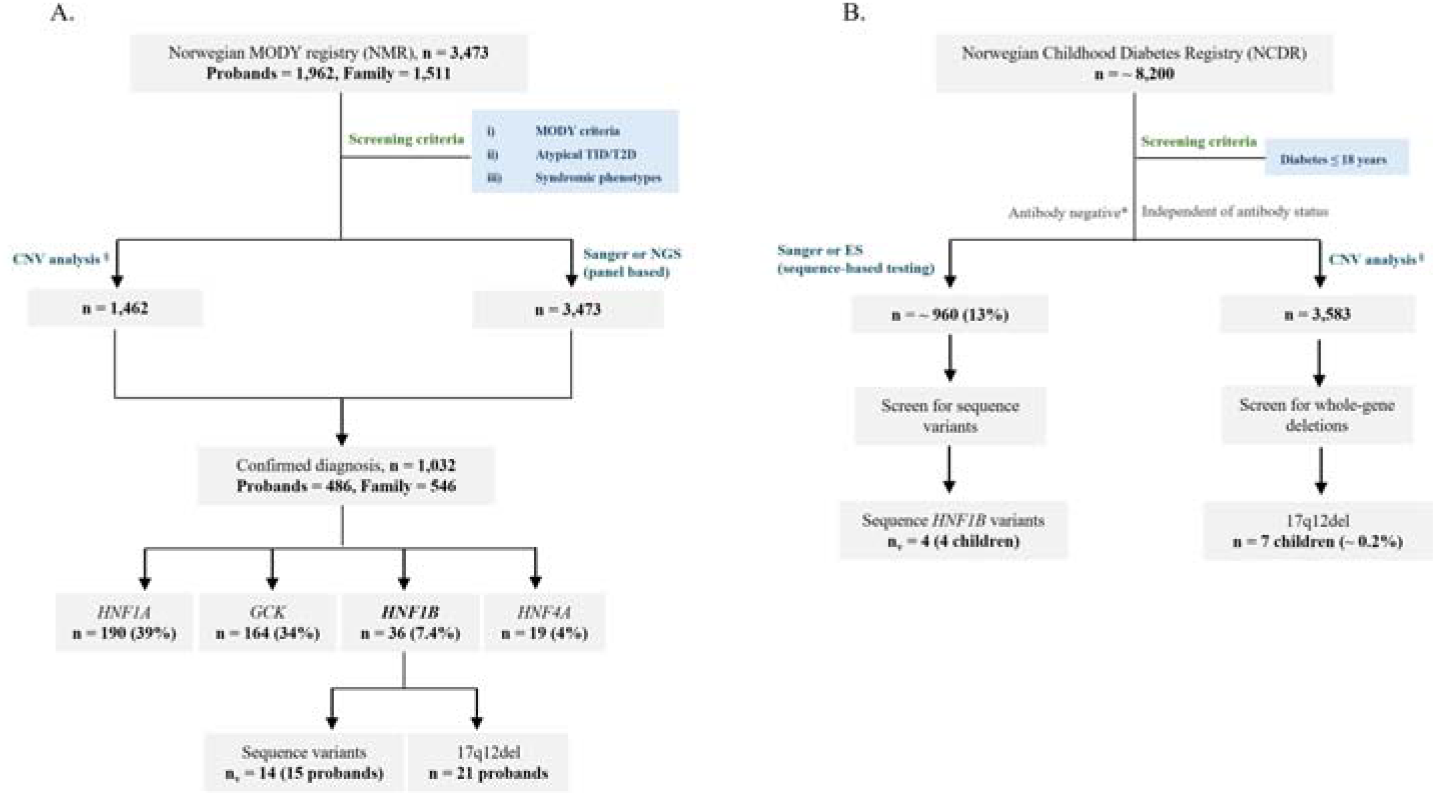
Workflow from referral to genetic diagnosis in the NMR (A) and NCDR (B). In Panels A and B, “n” denotes the number of individuals unless otherwise specified, whereas “n_v_” indicates the number of unique sequence variants. Percentages in Panel A were calculated based on the number of probands (n = 486) genetically diagnosed with monogenic diabetes. Approximately 87% of individuals in the NCDR were autoantibody-positive at diagnosis and are therefore not routinely screened for monogenic diabetes; however, ^*^469 age-and sex-matched autoantibody-positive children were included in the genetic analyses as controls, as detailed in[1]. ^§^CNV analysis was performed in a subset of individuals from the NMR and NCDR based on DNA availability and study design, as described in[14]. T1D = type 1 diabetes. T2D = type 2 diabetes. CNV = copy-number variant. 17q12del = 17q12 deletion. ES = exome sequencing. NGS = next-generation sequencing.

**Figure 2.**
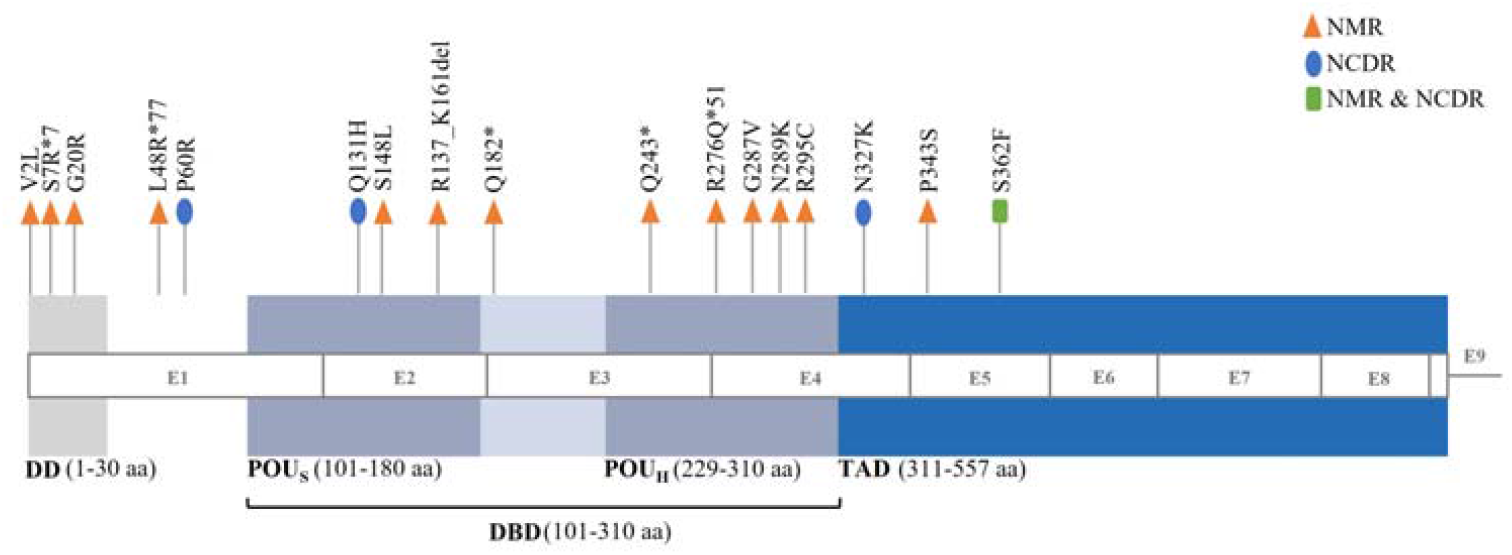
HNF-1 B protein domains and sequence variants identified in Norwegian registries. HNF-1B protein consists of an N-terminal dimerization domain (DD), conserved DNA-binding domain (DBD), and a disordered transactivation domain (TAD). Color coding distinguishes different functional domains, and the corresponding size of each domain is described by amino acids (aa), and each variant is labeled according to its respective position. Variants from the NMR and NCDR registries were color-coded to distinguish between cohorts. E = Exons. aa = amino acids. POU = Pit-1, Oct-1/2, UNC-86. POU_S_ = POU-specific domain; POU_H_ = POU homeodomain.

### 2.2 Clinical characteristics of LP/P sequence variants and 17q12del carriers

Clinical characteristics of individuals carrying LP/P sequence variants and 17q12del are summarized in Table 1. Renal involvement was universal in both groups, confirming renal disease as the core clinical feature of *HNF1B*-associated disease. Structural renal anomalies were more consistently observed among 17q12del carriers, whereas sequence variant carriers demonstrated greater phenotypic variability. Diabetes was common but not universal in both groups. While three individuals presented with isolated diabetes (e.g. carriers of the p.G20R variant), renal and extra-renal manifestations were frequently observed in the absence of diabetes, underscoring that diabetes is not required for clinical suspicion of *HNF1B*-disease.

Beyond diabetes and renal abnormalities, 17q12del patients also presented with neurocognitive impairment and electrolyte disturbances (hypomagnesemia). The apparent absence of these features among LP/P sequence variant carriers and other 17q12del carriers may reflect a lack of systematic screening rather than true absence of the phenotype. Pancreatic, genital tract, and liver abnormalities were also observed in both groups. Overall, clinical burden assessed using the Faguer score[15], was comparable between both 17q12del and LP/P sequence variant carriers. Family pedigrees of probands carrying *HNF1B* sequence variants are shown in Figure 3.

**Figure 3.** Pedigrees of probands in the NMR carrying *HNF1B* sequence variants. This figure has been removed. Please, contact the corresponding author (P.R.N.) to request access to this information.

### 2.3 Registry-level prevalence of *HNF1B* alterations

Seven children (approximately 0.2%) in the NCDR were diagnosed with HNF1B-MODY due to 17q12del. Among the 3,583 children screened for 17q12del, the prevalence is 0.195% (95% CI 0.09–0.40%), corresponding to 1.95 per 1,000 children (95% CI 0.95–4.03). When considering the entire NCDR cohort with approximately 8,200 children, this corresponds to a lower-bound prevalence estimate of 85 per 100,000 (95% CI 41-176). Of these seven children, three (43%) were positive for pancreatic autoantibodies at diagnosis. One child demonstrated elevated glutamic acid decarboxylase antibodies (GADA; 0.41 antibody index [AI], cut-off < 0.09 AI), while two children showed elevated insulinoma-associated antigen-2 antibodies (IA-2A; > 3.00 AI and 0.17 AI, cut-off < 0.11 AI, respectively). All three patients demonstrated a clinical and biochemical phenotype suggesting autoimmune diabetes with no or low insulin production. Among 486 probands in NMR with genetically confirmed monogenic diabetes, HNF1B-MODY accounted for 7.40% (95% CI 5.39-10.08%). When stratified by mutational mechanism, 17q12del were identified in 21/1,462 probands (1.43%, 95% CI 0.94-2.18%), while LP/P sequence variants were detected in 15/1,962 probands (0.76%, 95% CI 0.46-1.25%). Notably, one 17q12del carrier in the NMR cohort demonstrated clear positivity for all three autoantibodies (GADA 14.4 U/ml; cut-off < 5.0 U/ml, IA-2A 237 U/ml; cut-off < 7.5 U/ml, and zinc transporter 8 antibodies (ZnT8A) 33.6 U/ml; cut-off < 15 U/ml), consistent with an overlap with type 1 diabetes and a diabetes phenotype primarily due to type 1 diabetes, with an additional phenotype attributed to the 17q12del.

### 2.4 Functional analyses

In this study, functional impact of variants was examined on the full-length HNF-1B using *in vitro* protein assays. Functionally investigated variants include ten missense (p.V2M, p.G20R, p.P60R, p.Q131H, p.G287V, p.N289K, p.R295C, p.N327K, p.P343S, and p.S362F), three frameshift (p.S7R*7, p.L48R*77, and p.R276Q*51), two nonsense (p.Q182* and p.Q243*), and one in-frame deletion (p.R137_K161del) variant (Figure 2). To aid interpretation, five variants were incorporated as controls: two well-established pathogenic variants (p.S148L and p.R177*[16]), and two synonymous variants (p.T186= and p.V413=) without predicted splice effects (spliceailookup.broadinstitute.org). The p.V413= variant is observed in population data (gnomAD v4.1.0 Grpmax FAF = 0.006%).

#### 2.4.1 *HNF1B* gene variants and their classification prior to functional investigations

Among 17 identified sequence variants, ten (p.V2L, p.S7R*7, p.L48R*77, p.P60R, p.Q131H, p.Q243*, p.R276Q*51, p.N327K, p.P343S, and p.S362F) have not been previously reported. Five (p.G20R (personal communication and ClinVar VCV000591835.2)[17], p.Q182*[18], p.S148L[10, 18-23], p.G287V (ClinVar VCV000591835.2), p.N289K (personal communication), p.R295C[8, 24-27]) were reported by others or listed in ClinVar (www.ncbi.nlm.nih.gov/clinvar). The p.R137_K161del variant was previously reported and functionally characterized by our group[28]. Prior to functional analyses, all variants were classified according to American College of Medical Genetics and Genomics and the Association for Molecular Pathology (ACMG–AMP) guidelines, with specifications from the Clinical Genome Resource (ClinGen) (Table 2). Protein-truncating variants (PTVs) were classified as LP/P consistent with a LoF mechanism[29]. The p.R137_K161del variant was LP, supported by segregation with *HNF1B*-related disease[28]. Among the missense variants, five (p.G20R, p.S148L, p.G287V, p.N289K, and p.R295C) had sufficient non-functional evidence from previous studies, and/or additional clinical and family data from this cohort, enabling variants to be classified as LP/P. Irrespective of the variant class, all identified variants were incorporated in the functional analyses.

**Table 2.**
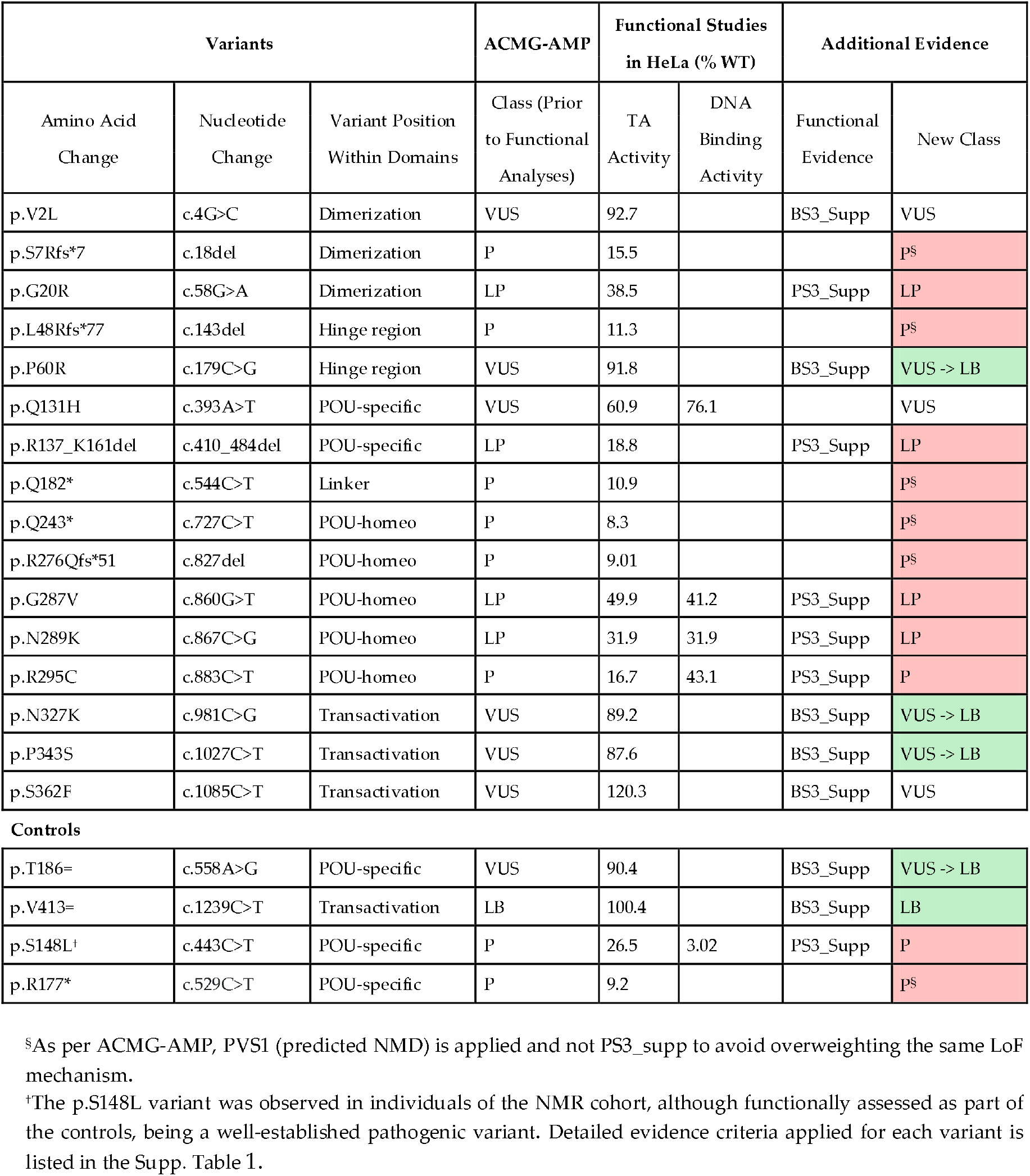
ACMG-AMP variant classification before and after functional analysis.

| Variants |  |  | ACMG-AMP | Functional Studies<br>in HeLa (% WT) |  | Additional Evidence |  |
| --- | --- | --- | --- | --- | --- | --- | --- |
| Amino Acid<br>Change | Nucleotide<br>Change | Variant Position<br>Within Domains | Class (Prior<br>to Functional<br>Analyses) | TA<br>Activity | DNA<br>Binding<br>Activity | Functional<br>Evidence | New Class |
| p.V2L | c.4G>C | Dimerization | VUS | 92.7 |  | BS3_Supp | VUS |
| p.S7Rfs*7 | c.18del | Dimerization | P | 15.5 |  |  | P<sup>§</sup> |
| p.G20R | c.58G>A | Dimerization | LP | 38.5 |  | PS3_Supp | LP |
| p.L48Rfs*77 | c.143del | Hinge region | P | 11.3 |  |  | P<sup>§</sup> |
| p.P60R | c.179C>G | Hinge region | VUS | 91.8 |  | BS3_Supp | VUS-> LB |
| p.Q131H | c.393A>T | POU-specific | VUS | 60.9 | 76.1 |  | VUS |
| p.R137_K161del | c.410_484del | POU-specific | LP | 18.8 |  | PS3_Supp | LP |
| p.Q182* | c.544C>T | Linker | P | 10.9 |  |  | P<sup>§</sup> |
| p.Q243* | c.727C>T | POU-homeo | P | 8.3 |  |  | P<sup>§</sup> |
| p.R276Qfs*51 | c.827del | POU-homeo | P | 9.01 |  |  | P<sup>§</sup> |
| p.G287V | c.860G>T | POU-homeo | LP | 49.9 | 41.2 | PS3_Supp | LP |
| p.N289K | c.867C>G | POU-homeo | LP | 31.9 | 31.9 | PS3_Supp | LP |
| p.R295C | c.883C>T | POU-homeo | P | 16.7 | 43.1 | PS3_Supp | P |
| p.N327K | c.981C>G | Transactivation | VUS | 89.2 |  | BS3_Supp | VUS-> LB |
| p.P343S | c.1027C>T | Transactivation | VUS | 87.6 |  | BS3_Supp | VUS-> LB |
| p.S362F | c.1085C>T | Transactivation | VUS | 120.3 |  | BS3_Supp | VUS |
| **Controls** |  |  |  |  |  |  |  |
| p.T186= | c.558A>G | POU-specific | VUS | 90.4 |  | BS3_Supp | VUS-> LB |
| p.V413= | c.1239C>T | Transactivation | LB | 100.4 |  | BS3_Supp | LB |
| p.S148L<sup>†</sup> | c.443C>T | POU-specific | P | 26.5 | 3.02 | PS3_Supp | P |
| p.R177* | c.529C>T | POU-specific | P | 9.2 |  |  | P<sup>§</sup> |

<sup>§</sup>As per ACMG-AMP, PVS1 (predicted NMD) is applied and not PS3\_supp to avoid overweighting the same LoF mechanism.
<sup>†</sup>The p.S148L variant was observed in individuals of the NMR cohort, although functionally assessed as part of the controls, being a well-established pathogenic variant. Detailed evidence criteria applied for each variant is listed in the Supp. Table 1.

ACMG-AMP-ClinGen guidelines without functional evidence were used to assign an initial classification. Transactivation and DNA-binding activities of variants were presented as % of WT. Functional evidence (e.g., PS3_Supp or BS3_Supp) was added to the classification based on the obtained transactivation data in HeLa cells. Variants with impaired transactivation activity (≤ 50%) were assigned PS3_Supp, while variants with comparable activity to WT HNF-1B (≥ 85%) were assigned BS3_Supp, in accordance with the Brnich control criteria[30]. Variants that were reclassified after functional analyses are highlighted with an arrow. Final classifications are coloured in green for likely benign (LB) and red for likely pathogenic/pathogenic (LP/P) variants. POU = Pit-1, Oct-1/2, UNC-86. Supp = Supporting. VUS = variants of uncertain significance.

#### 2.4.2 Effect of *HNF1B* variants on transactivation activity

Transactivation potential of *HNF1B* variants examined in lysates of transfected HeLa and MIN6 cell lines revealed similar profiles across both cell lines (Figure 4). All PTVs (p.S7R*7, p.L48R*77, p.Q182*, p.Q243*, and p.R276Q*51) and the deletion variant (p.R137_K161del) showed markedly reduced activity (≤ 20% in HeLa and ≤ 30% in MIN6) similar to HNF1B-MODY controls. Four missense variants (p.G20R, p.G287V, p.N289K, and p.R295C) located in either DD or DBD, also showed significantly impaired transactivation potential (≤ 50% in HeLa cells and ≤ 55% in MIN6 cells). The p.Q131H variant demonstrated, moderately reduced activity (60% in HeLa cells and 80% in MIN6 cells). All other variants demonstrated transcriptional activity comparable to WT HNF-1B (Figure 4).

**Figure 4.**
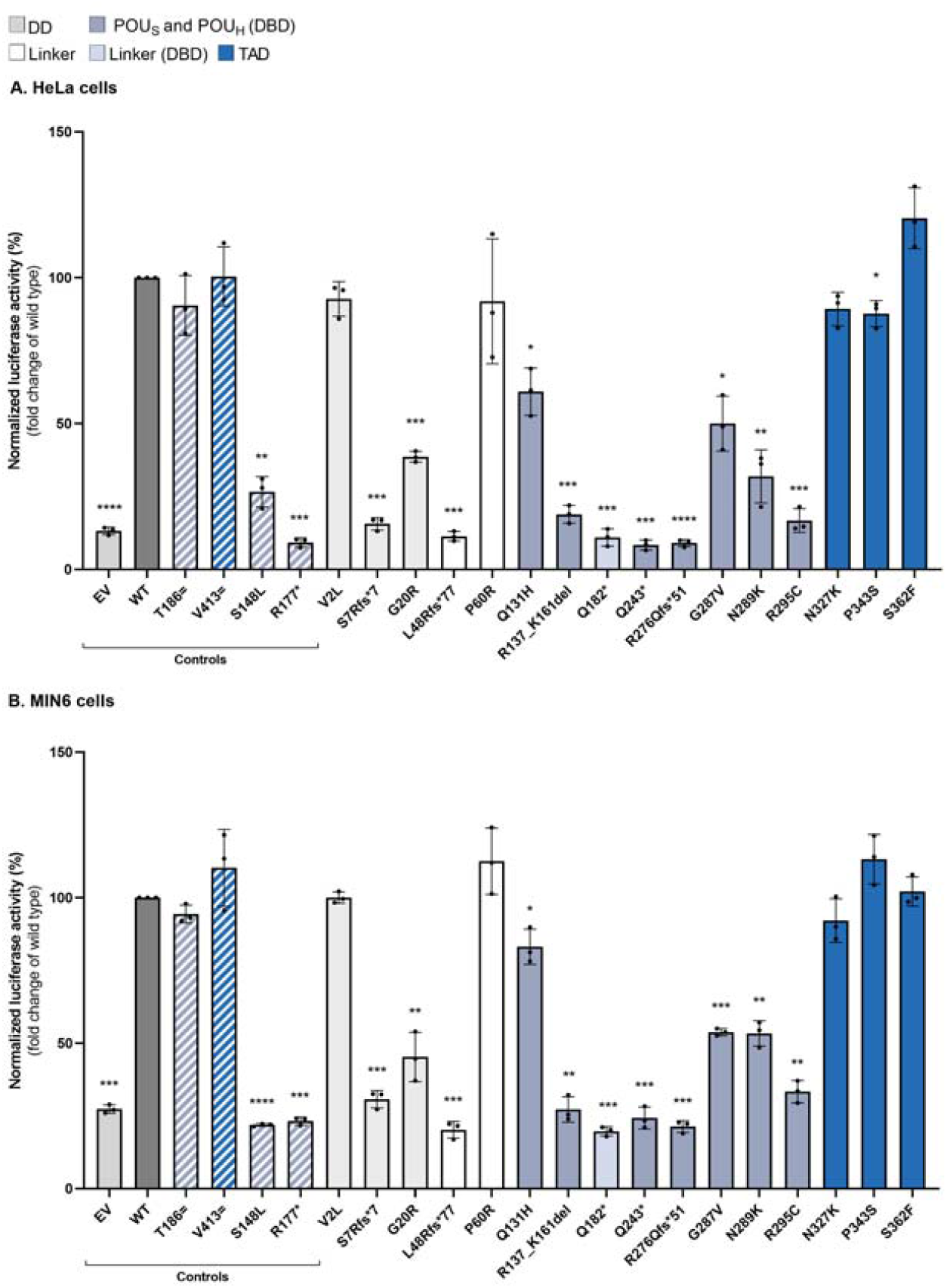
Transactivation profile of HNF-1B protein variants in HeLa (A) and MIN6 (B) cells. Expression of luciferase was measured after co-transfecting cells with reporter plasmids encoding firefly luciferase (pGL3-RA for HeLa and pGL3-GLUT2 for MIN6), Renilla luciferase (pRL-SV40) alongside EV, WT or HNF1B variant plasmids. The cells were lysed 24-(HeLa) or 48-hour (MIN6) post-transfection, followed by measurements of transactivation activities. The relative transactivation activity of each variant was expressed as a percentage of WT activity (set to 100%). Each bar represents the mean transactivation activity of all three biological replicates, with three dots representing three individual runs. Standard deviations were denoted by error bars and the asterisks (*) represent statistical significance levels (*p ≤ 0.05; **p ≤ 0.01; ***p ≤ 0.001; ****p ≤ 0.0001). EV = empty vector, WT = wild type.

#### 2.4.3 Nuclear import and DNA-binding analysis of *HNF1B* variants

The deletion variant and missense variants in the DBD with impaired transactivation activity were evaluated for their ability to bind to the rat albumin (RA) promoter using electrophoretic mobility shift assays (EMSA). PTVs were excluded from this analysis due to the expected nonsense-mediated decay (NMD) and/or loss of one or more protein domains (DD, DBD and/or TAD) in the unlikely event of NMD-escape. Among the assessed missense variants (p.Q131H, p.G287V, p.N289K, and p.R295C), all except p.Q131H showed markedly impaired DNA binding (≤ 45% activity vs. the WT) in both cell lines. The p.Q131H variant exhibited a modest reduction in DNA binding (∼ 75% activity), whereas the deletion variant p.R137_K161del showed minimal to no DNA binding in EMSA, likely due to markedly reduced protein expression in combination with impaired DNA-binding capacity (Figure 5 and Supp. Figure 1).

**Figure 5.**
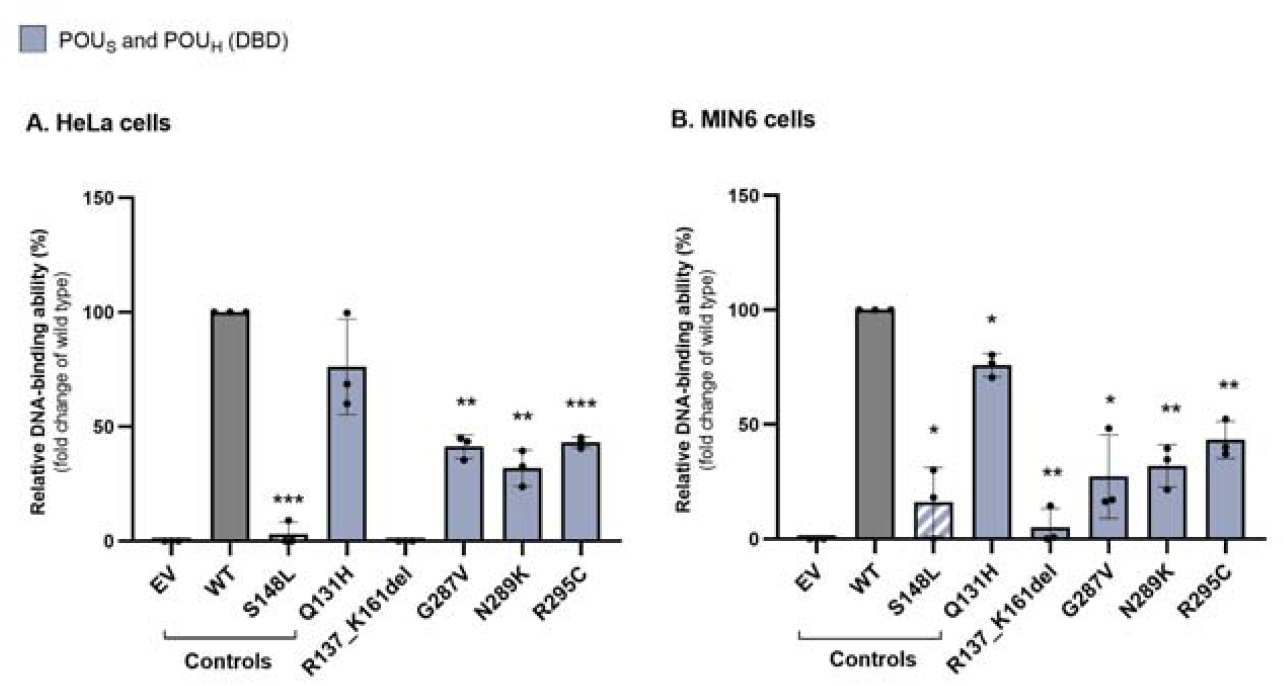
DNA binding capabilities of HNF-1B protein variants from transfected HeLa (A) and MIN6 (B) cells. HeLa and MIN6 cells transfected with EV, HNF1B WT, or variant plasmids were lysed, fractionated, and obtained nuclear lysates were used to test DNA binding abilities. Equal amounts (5 µg in HeLa cells and 10 µg in MIN6 cells) of nuclear fractions were incubated with a Cy5-labeled oligonucleotide containing the HNF-1B binding site in the RA-promoter. HNF-1B-DNA complexes were separated on a 6% retardation gel, and the fluorescence signal was quantified. The relative DNA binding capacity of each variant was expressed as a percentage of WT activity (set to 100%). The average DNA binding activity of all three biological replicates was shown in each bar, with three dots representing three individual replicates. The standard deviations were denoted by error bars and the asterisks (*) represent statistical significance levels (*p ≤ 0.05; **p ≤ 0.01; ***p ≤ 0.001). EV = empty vector, WT = wild type.

### 2.5 Functional and clinical correlations

To assess concordance between functional activity and clinical severity, we analyzed transactivation activity (%) versus Faguer score across 17 variants. Spearman’s rank correlation showed significant inverse association (ϱ = –0.701, p = 0.002; 95% CI –0.887 to –0.318), indicating that lower transactivation level corresponds to greater clinical severity. Except for p.G20R, all variants with impaired transactivation activity exhibited Faguer scores of 8 or higher (Figure 6).

**Figure 6.**
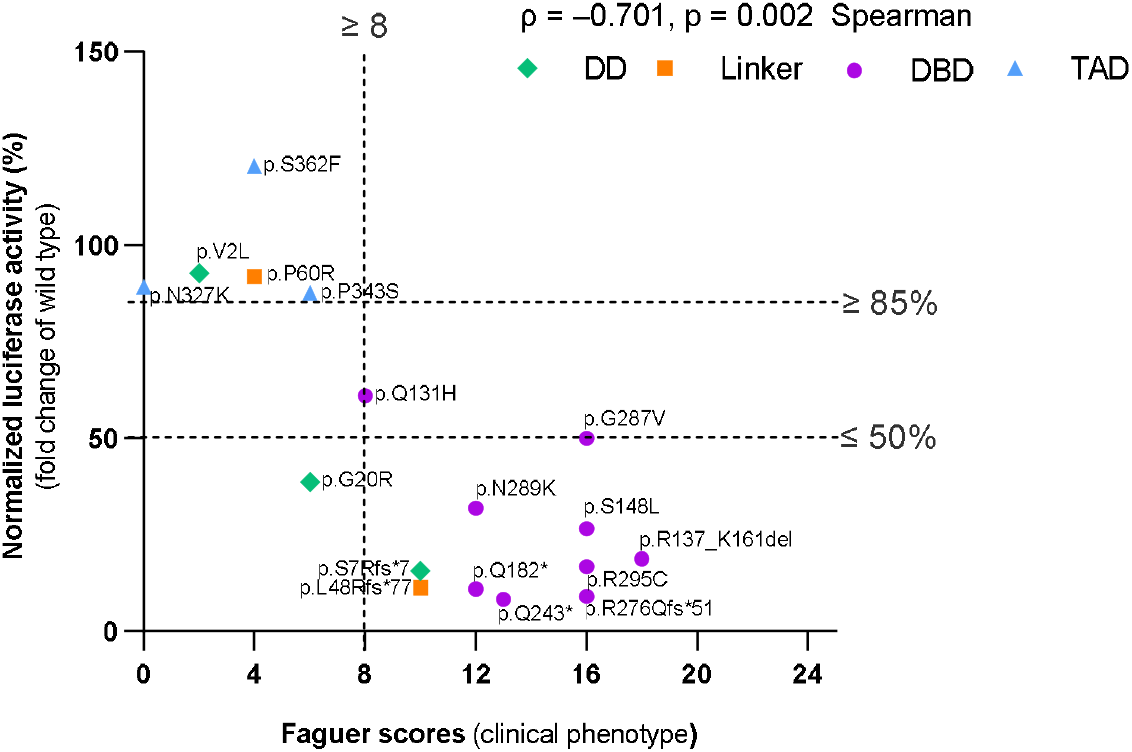
Correlation between transactivation activity and Faguer scores. Transactivation capacity of variants assessed in HeLa cells (expressed as a percentage relative to wild type, 100%) is plotted on the y-axis, while the highest Faguer scores obtained for each variant, representing clinical phenotype severity are shown on the x-axis. Each data point represents a variant colored by its protein domain: green for dimerization domain (DD; residues 1–30), orange for linker region (31–100), red for DNA-binding domain (DBD; 101–310), and blue for transactivation domain (TAD; 311–557). The vertical dashed line represents a Faguer score of 8 set as a threshold for applying the PP4 criteria. The horizontal lines at 50 and 85% were included as a visual reference for impaired and normal transactivation activity, respectively.

To further evaluate the clinical relevance of transactivation activity, variants were divided into two groups based on activity, ≤ 50% vs. > 50% of WT. Variants with ≤ 50% transactivation (n = 11) had a significantly higher median Faguer score (median = 13.0, interquartile range [IQR] = 11-16), compared to variants in the > 50% activity group (n = 6), with a median score of 4.0 (IQR = 2-6). The difference was statistically significant (U = 1.5, Z = −3.193, exact p < 0.001). The mean Faguer score in the ≤ 50% group was 13 ± 3.6, compared to 4.0 ± 2.8 in the > 50% group. These findings underscore that the PTVs and DD/DBD missense variants with ≤ 50% transactivation activity are functionally deleterious and are associated with renal anomalies and/or extra-renal features, while missenses in the linker/TAD regions (e.g., p.V2L, p.P60R, p.N327K, p.P343S, and p.S362F) retained normal HNF-1B function with no specific *HNF1B* abnormalities. There were no significant differences in Faguer score severity between protein truncating and damaging missense variant carriers, although this may partly be due to the small sample size.

## Discussion

In this study, we have addressed gaps in *HNF1B* variant interpretation by functionally testing variants identified in patients from two Norwegian diabetes registries. While *in vitro* assays cannot capture all pathogenic mechanisms, functional results generated using our cDNA system aligned with clinical phenotype (Faguer scores) of carriers, supporting causality of 11 LP/P variants. Functional data exclusively enabled reclassification of three variants (p.P60R, p.N327K, and p.P343S) to LB and provided data to reduce the likelihood of pathogenicity of two VUS (p.V2L and p.S362F). The finding of impaired transactivation activity in functional assays for two variants in the NMR initially classified as VUS (p.G287V and p.N289K) prompted targeted clinical and segregation follow-up, which subsequently provided additional evidence enabling reclassification to LP.

Across cohorts, *HNF1B* accounted for 7.4% of monogenic diabetes cases in the NMR (among 486 probands with genetically confirmed MODY) and ∼ 0.2% in the NCDR (among 3,583 individuals screened). Since CNV analysis was not performed systematically in all individuals, 17q12del may theoretically have been missed in a subset of individuals in both registries. In total, across both registries, 26 children aged 0-18 years at the time of diagnosis were identified as having HNF1B-MODY. Within the NMR cohort, pediatric carriers of the 17q12del were primarily ascertained based on structural renal anomalies rather than the clinical presentation of diabetes. Notably, autoantibody positivity did not exclude HNF1B-MODY, as three children with the 17q12del were positive for GADA and/or IA-2 antibodies. Clinically, these findings support genetic testing for *HNF1B* in children with *HNF1B*-consistent features, irrespective of diabetes or single autoantibody positivity, to reduce missed diagnoses and enable early detection and management of associated comorbidities.

### Variants with (likely) pathogenic effects

The p.G20R variant was identified in two unrelated families within the NMR. In the F4 pedigree, the proband, a sibling, and their father carried the variant; all developed diabetes, and the father had gout, a feature compatible with HNF1B-MODY[31]. Recurrence of the variant across multiple unrelated families (ClinVar VCV000591835.2; accessed January 19, 2026), its segregation through three meiosis[17] and the markedly reduced transactivation activity together support a LP classification (Figure 4). Notably, none of the p.G20R carriers in our cohort had known renal disease or structural renal abnormalities at diagnosis, underscoring the need for clinical surveillance in families (Table 2).

Four missense variants in the DBD (p.S148L, p.G287V, p.N289K, and p.R295C) were classified as LP/P. All these variant carriers reported renal, pancreatic anomalies and/or diabetes. In family F18, the proband harboring the p.R295C variant has renal failure. Moreover, this residue has repeatedly been linked to renal and extra-renal involvement, warranting close surveillance of the family members in our cohort[8, 20, 26]. Besides, alternative variants of N289 and R295 residues (e.g., p.N289D (unpublished data), p.R295P, p.R295H) have also been reported to cause HNF1B-MODY[10, 19, 20]. The p.R137_K161del (24 aa deletion) classified as LP was identified in five related individuals, all with diabetes, renal and extra-renal features[28].

Three frameshift (p.S7R*7, p.L48R*77, and p.R276Q*51) and two nonsense (p.Q182* and p.Q243*) variants were classified as LP/P according to ACMG-AMP and demonstrated consistent LoF in our cDNA-based assays (Figure 4). Based on the position of the premature stop codons, these variants are predicted to trigger NMD of the mRNA transcript; however, even in the unlikely event that they escape NMD, they are still expected to produce truncated proteins lacking key functional domains. Because the expression plasmids employed in functional assays lack intronic sequences, transcripts of these protein truncating variants may escape NMD *in vitro*. As a result, these assays do not recapitulate the physiological mRNA degradation process but instead assess the residual activity of any truncated protein that is produced.

### (Likely) benign variants

Functional and clinical evidence supported reclassification of p.P60R, p.N327K, and p.P343S variants to LB (Table 2). Population frequency data met BS1 for p.P60R (Grpmax FAF = 0.0083%) and p.N327K (Grpmax FAF = 0.0055%; gnomad.broadinstitute.org; accessed January 19, 2026) according to *HNF1A* frequency thresholds (clinicalgenome.org/affiliation/50016). Phenotypically, the p.P60R carrier was positive for GADA (0.12 AI; cut-off < 0.09 AI). The p.N327K carrier was positive for both GADA (1.54 AI; cut-off < 0.09 AI) and IA-2A (0.38 AI; cut-off < 0.11 AI) and with markedly reduced C-peptide levels (< 50 pmol/L; reference 220–1,400 pmol/L), consistent with autoimmune type 1 diabetes. The p.P343S proband was obese (BMI 37.9 kg/m^2^) with high C-peptide levels (2360 pmol/L), suggestive of type 2 diabetes/insulin resistance. A family history of diabetes was absent for p.P60R and p.N327K variant carriers; for p.P343S, the mother was also a carrier, but available data did not demonstrate a segregating *HNF1B* phenotype.

### Variants with uncertain significance

Two variants (p.V2L and p.S362F) remained VUS (Table 2) after functional evaluation. Both showed normal transactivation (≥ 85%) in HeLa and MIN6 cells. Clinically, carriers had diabetes without extra-pancreatic features. The p.V2L carrier was diagnosed at an age range of 35-40 years with elevated C-peptide levels (2,220 pmol/L), suggestive of insulin resistance; and the p.S362F proband was obese (BMI 30.1 kg/m^2^). Transactivation activity levels comparable with WT (PS3_Supporting) for these variants and the absence of *HNF1B*-specific features argue against *HNF1B* causality. Alternative etiologies such as type 2 diabetes remain possible. For the last variant, p.Q131H, limited clinical data and modest functional impairment (Figure 4 and 5) provided insufficient evidence to classify it as either LB/B or LP/P; thus, it remains a VUS.

## 4. Materials and Methods

### 2.1 Study participants and genetic testing

The NMR is a nationwide, population-based registry established in 1997[21]. The registry comprises established and suspected cases of monogenic diabetes referred by physicians for genetic testing. By April 2025, NMR comprised 3,473 participants (1,962 probands and 1,511 family members) and the registry was established for diagnostic and research purposes, with the goal of improving diagnosis and treatment of these monogenic disorders. In routine practice, clinicians refer patients based on a clinical suspicion of monogenic diabetes using following criteria: (1) diabetes in a first-degree relative; (2) onset before ∼ 35 years of age in at least one family member; (3) absence of pancreatic autoantibodies; (4) BMI ≤ 30 kg/m^2^, and (5) atypical presentation of type 1 diabetes (low insulin requirements (< 0.5 U/kg/day), no autoantibodies, or an unusual disease course)[21, 32]. However, a patient does not necessarily need to meet all criteria for referral and subsequent analysis. Analyses of diabetes-related autoantibodies were performed at two different laboratories (the Hormone Laboratory at Haukeland University Hospital and Aker/Oslo University Hospital), using different methods and laboratory-specific cut-off values. Genomic DNA was extracted using standard procedures. Patients with suspected monogenic diabetes underwent genetic testing: prior to 2020, clinical judgement-directed Sanger sequencing of *HNF1A, GCK, HNF1B, HNF4A, INS, KCNJ11*, and *ABCC8*; from 2020 onward, panel-based exome sequencing.

The NCDR includes approximately 8,200 participants (July 2023) diagnosed with diabetes before 18 years of age since 2002[3], covering ∼ 99.6% of pediatric cases in Norway. At diagnosis, clinical data, biochemical measures (C-peptide, GADA, IA-2A, and ZnT8A), DNA, and serum are collected. In total, 960 antibody negative individuals (13% of all participants) as well as 469 age-and sex-matched participants with autoimmune diabetes were investigated for variants in monogenic diabetes genes (including *HNF1B*) using panel sequencing or exome sequencing[1, 2]. In addition, 1,462 individuals included in the NMR and 3,583 of the ∼ 8,200 individuals included in the NCDR were additionally genotyped using the Illumina Infinium Global Screen Array-24 v2.0 to detect whole-gene deletions and Multiplex Ligation-Dependent Probe Amplification (MLPA) was subsequently used to confirm pathogenic CNVs[14]. However, in the NMR, MLPA-based CNV analysis was implemented routinely in parallel with panel-based testing from 2016 onwards. Prior to 2016, CNV testing was not performed systematically, and approximately 200 probands without a confirmed genetic diagnosis were not assessed for CNVs. Figure 1 presents the stepwise flow chart from referral to genetic testing, and subsequent genetic diagnosis in the NMR/NCDR registries.

### 2.3 Clinical data

Clinical characteristics of 17q12del and LP/P sequence variant carriers are summarized in Table 1. Continuous variables include age at referral, age at diabetes diagnosis, BMI, and HbA1c. Categorical variables include sex, registry source (NMR/NCDR), diabetes at diagnosis, insulin at referral, and established HNF1B-MODY features.

### 2.4 *HNF1B* constructs and cell culture

For plasmid construction, variants were introduced into the full-length human *HNF1B* cDNA (NCBI NM_000458.3) within the pcDNA3.1/His B vector via the QuikChange II XL Site-Directed Mutagenesis Kit (Agilent Technologies #200522-5, Santa Clara, CA, USA) utilizing the primer sets detailed in Supp Table 2. Plasmids were transformed into ultracompetent E. coli cells (DH5α) and DNA was extracted using the plasmid midi kit (QIAGEN #12243, Hilden, Germany). Introduction of the genetic variant was verified by Sanger sequencing using BigDye Terminator v.3.1 on an ABI 3100 Genetic Analyzer. *In vitro* assessments were performed in HeLa (CCL-2 ATCC, Manassas, VA, USA) and MIN6 cells, received from Prof. RN Kulkarni at Joslin Diabetes Center (Harvard Medical School, Boston, MA, USA). While the former is derived from human cervical epithelial tissue, the latter is a pancreatic beta cell line originating from transgenic C57BL/6 mouse insulinoma. Importantly, HeLa cells do not express HNF-1B endogenously, whereas MIN6 allows for the investigation of *HNF1B* variants under more physiologically suitable conditions. Both cell types were cultured, maintained, and transiently transfected using Lipofectamine 2000 Transfection Reagent (Invitrogen, Thermo Fisher Scientific #11668-019, Waltham, MA, USA) and Opti-MEM Reduced Serum Medium (Gibco, Thermo Fisher Scientific #31985-062, Waltham, MA, USA). A commercial vector encoding green fluorescent protein (GFP) (Amaxa, Lonza #V4XC-2024, Cologne, Germany) served as a control for transfection efficiency.

### 2.5 Protein functional analyses

Functional analyses were performed to assess the transactivation activity, DNA-binding and subcellular localization of variants in HeLa and MIN6 cells as described in our previous study[33].

#### 2.5.1 Transactivation assay

To assess the transactivation potential of variants, HeLa and MIN6 cells were transiently transfected with individual plasmids containing *HNF1B* variants, in addition to an EV and WT *HNF1B* cDNA, and a total of four control variants. Reporter vectors containing the firefly luciferase gene fused to either a rat albumin (RA) or glucose transporter 2 (*GLUT2*) gene promoter, as well as an internal control vector with the constitutively active simian virus 40 (SV40) promoter linked to the *Renilla* luciferase gene, were co-transfected into the cells. After 24 or 48 hours, the cells were harvested, and the luciferase activities measured using the Dual-Luciferase Reporter Assay System (Promega #E1910, Fitchburg, WI, USA) with a Centro XS3 LB 960 microplate luminometer (Berthold Technologies). The functional effect of each variant was estimated relative to WT HNF-1B activity after normalization of firefly luciferase against *Renilla* luciferase activity.

#### 2.5.2 Subcellular fractionation

Subcellular fractionation was performed in order to isolate nuclear and cytosolic cell fractions from transiently transfected HeLa and MIN6 cells. In short, pellets from whole-cell lysates were first suspended in a buffer (A) consisting of 10 mM HEPES (pH 7.8), 1.5 mM MgCl_2_, 10 mM KCl, 0.10% (v/v) IGEPAL, 0.5 mM DTT, and 1 cOmplete Mini EDTA-free Protease Inhibitor Cocktail tablet (Sigma-Aldrich #11836170001, St. Louis, MO, USA), with the total volume adjusted to 10 ml with Milli-Q water. Following incubation and brief centrifugation at 16,250 x g (4°C), the supernatants, containing the cytosolic fractions, were collected. Next, the pellets were washed twice in buffer A before thorough resuspension in a second buffer (B) comprised of 20 mM HEPES (pH 7.8), 420 mM NaCl, 1.5 mM MgCl_2_, 0.2 mM EDTA, 0.5 mM DTT, and 1 cOmplete Mini EDTA-free Protease Inhibitor Cocktail tablet (Sigma-Aldrich #11836170001), again with the overall volume adjusted to 10 ml with Milli-Q water. The pellets were then vortexed and finally centrifuged for 15 minutes at 16,250 x g (4°C) before harvesting the nuclear fractions.

Prior to SDS-PAGE and immunoblotting, the total amount of protein in each sample was quantitated using the Pierce BCA Protein Assay Kit (Thermo Fisher #23225 Waltham, MA, USA), ensuring an equal loading amount of 5 µg protein from each sample. In addition to anti-HNF-1B antibody (Sigma-Aldrich #HPA002083), antibodies against topoisomerase I (Abcam #ab196642, Cambridge, UK) and topoisomerase II alpha (Cell Signaling Technology #12286S Danvers, MA, USA) served as nuclear markers for HeLa and MIN6 cells, respectively, and alpha-tubulin (Abcam #ab40742) served as a cytosolic marker in both cell lines. *HNF1B* protein levels were quantified by normalizing to topoisomerase I or II alpha to obtain HNF-1B/topoisomerase ratios. These ratios were expressed relative to WT and used to adjust the amount of nuclear protein required for EMSA.

#### 2.5.3 DNA-binding studies

The DNA-binding abilities of the five variants situated in the DBD of the HNF-1B protein were studied using EMSA. In short, equal protein amounts of nuclear fractions from both HeLa (5 µg) and MIN6 cells (10 µg) transfected with these variants were incubated together with double-stranded cyanine 5 (Cy5)-labeled oligonucleotides containing the RA promoter (5’-TGT GGT TAA TGA TCT ACA GTT A-3’). Optimal binding conditions were facilitated using the Odyssey EMSA Buffer Kit (LI-COR Biosciences #829-07910, Lincoln, NE, USA), and the resulting reactions were separated by gel electrophoresis, visualized with the ChemiDoc MP imaging system (Bio-Rad Laboratories), and quantitated in terms of signal strength relative to WT HNF-1B.

### 2.6 Variant interpretation

HNF1B-specific variant interpretation guidelines have so far not been published by the ClinGen Monogenic Diabetes Expert Panel (MDEP). Thus, all variants were classified according to MDEP HNF1A-specific ACMG-AMP guidelines (clinicalgenome.org/affiliation/50016), adapted for the *HNF1B* gene where applicable. The ACMG-AMP framework provides a structured approach to variant classification, categorizing variants as LB/B or LP/P based on weighted evidence[29], with updates made to the general guidelines made by the ClinGen[34]. Benign criteria applied in this study include BS1, BS3_Supporting, BP4, and BP7 while pathogenic criteria comprise PVS1, PS4/PS4_Moderate, PM1/PM1_Supporting, PM4_Moderate, PS2/PM6_Supporting, PS2/PM6_Moderate, PS2/PM6_Very strong, PP1/PP1_Moderate/PP1_Strong, PP3, PP4, PM2_Supporting, and PS3_Supporting. For individual criterion details and cut-offs, refer to Supp Table 3. AlphaMissense was used as an *in silico* prediction tool to support application of the BP4 and PP3 criteria[35]. Clinical features were scored according to the Faguer et al. specifications (PP4)[15]. PP4 criterion was applied if at least one of the variant carriers had a score of 8 (pre-specified threshold) or higher.

### 2.7 Statistics

Statistical analyses were performed using SPSS (version 29.0.2.0). Results are presented as mean with standard deviation or as medians with interquartile ranges, as appropriate. Continuous variables were summarized as median with 25th-75th percentiles, and categorical variables were summarized as counts and percentages (%). Non-parametric correlations were assessed using Spearman’s rank correlation coefficient (ϱ). Group comparisons of medians were performed using the Mann–Whitney U test. Statistical significance for transactivation and DNA-binding data was evaluated using two-tailed Student’s t-tests (Welch’s t-test) comparing each variant with WT (set as 100%). A p-value < 0.05 was considered statistically significant. Registry level prevalence was expressed as proportions with two-sided 95% confidence intervals (CIs) computed using the Wilson method. Functional and clinical data were plotted using GraphPad Prism 9 (version 9.5.1, GraphPad Software, San Diego, CA, USA).

## 5. Conclusions

Integration of functional assays with clinical and family data improved *HNF1B* variant interpretation and reduced diagnostic uncertainty in these registries. Importantly, *HNF1B* accounted for a significant proportion of MODY in our cohorts. Pediatric carriers of 17q12del were frequently identified through renal and extrapancreatic manifestations rather than diabetes alone, and positive autoantibody titers did not exclude HNF1B-MODY in children. When considering 17q12del and LP/P sequence variant carriers, we would miss at least 18 cases if using classical MODY criteria only, and three cases if not including classical MODY criteria. These findings indicate that HNF1B-MODY diagnosis is an exception among MODY subtypes, where reliance on classical MODY criteria may lead to missed diagnoses. Therefore, *HNF1B* genetic testing should continue to be considered in children with *HNF1B*-specific renal features irrespective of diabetes or antibody status. Early molecular diagnosis enables appropriate surveillance of renal and systemic complications, informs treatment decisions, and improves genetic counselling and long-term clinical management among carriers.

## Data Availability

All data produced in the present study are available upon reasonable request to the authors

## Supplementary Materials

The following supporting information is available at:

## Author Contributions

Conceptualization – P.S., L.B., J.V.S., B.B.J., J.M., and P.R.N.; Functional data curation – A.P., I.L.K., K.A.Mo., M.T., B.B.J., and J.M.; Formal analysis – A.P., E.T., P.S., I.L.K., K.A.Mo., M.T., B.B.J., and J.M.; Investigation – A.P., E.T., P.S., I.L.K., K.A.Mo., M.T., J.V.S., B.B.J., J.M., and P.R.N.; Methodology – A.P., E.T., I.L.K., K.A.Mo., M.T., C.B.-C., B.B.J., and J.M.; Supervision – J.V.S., B.B.J., J.M., and P.R.N.; Validation – A.P., E.T., P.S., I.L.K., K.A.Mo., M.T., B.B.J., and J.M.; Visualization – A.P., M.T.; Resources – B.B.J., J.M., and P.R.N.; Project administration – B.B.J., J.M., and P.R.N.; Writing – original draft – A.P.; Writing – review & editing – E.T., P.S., L.B., M.V., T.S., K.A.Ma, T.I.P., J.V.S., B.B.J., J.M., and P.R.N.; Funding acquisition – A.P., M.V., S.J., B.B.J., J.M., and P.R.N.

## Institutional Review Board Statement

The study was approved by the Regional Committee for Medical Research Ethics, Region West-Norway (Haukeland University Hospital, University of Bergen, Faculty of Medicine, PO Box 7804, NO-5020 Bergen, Norway) (project #18836 and #18848) and conducted in accordance with the Declaration of Helsinki. None other ethical oversight bodies have ruled over the study.

## Informed Consent Statement

Written informed consent was obtained from all participants or legal guardians of children under 16 years.

## Data Availability Statement

The data used in this study were obtained from Norwegian MODY and Norwegian Childhood Diabetes Registries and contain sensitive patient information. All original contributions presented in this study are included in the article/Supplementary Material. Detailed clinical tables can be provided upon request, and additional inquiries can be directed to the corresponding author.

## Acknowledgments

We gratefully acknowledge Liv Aasmul, Monika Ringdal, Louise Grevle, and Anne H. Knudsen for their help with laboratory logistics, plasmid preparation, procurement of reagents and primers, and technical support.

## Abbreviations

The following abbreviations are used in this manuscript:

MODY: Maturity-Onset Diabetes of the Young
NMR: Norwegian MODY Registry
NCDR: Norwegian Childhood Diabetes Registry
HNF-1B: Hepatocyte Nuclear Factor-1B
DD: Dimerization Domain
DBD: DNA-Binding Domain
TAD: Transactivation Domain
NLS: Nuclear Localization Signal
EV: Empty Vector
WT: Wild Type
EMSA: Electrophoretic Mobility Shift Assay
RA: Rat Albumin
GLUT2: Glucose Transporter 2
ACMG-AMP: American College of Medical Genetics and Genomics and the Association for Molecular Pathology
ClinGen: Clinical Genome
MDEP: Monogenic Diabetes Expert Panel
LP/P: Likely Pathogenic / Pathogenic
B/LB: Benign / Likely Benign
VUS: Variants of Uncertain Significance
NMD: Nonsense Mediated Decay

## References

1. Johansson, B.B., et al., Targeted next-generation sequencing reveals MODY in up to 6.5% of antibody-negative diabetes cases listed in the Norwegian Childhood Diabetes Registry. Diabetologia, 2017. 60(4): p. 625–635.

2. Shields, B.M., et al., Maturity-onset diabetes of the young (MODY): how many cases are we missing? Diabetologia, 2010. 53(12): p. 2504–8.

3. Svalastoga, P., et al., Characterisation of HNF1A variants in paediatric diabetes in Norway using functional and clinical investigations to unmask phenotype and monogenic diabetes. Diabetologia, 2023. 66(12): p. 2226–2237.

4. Zhang, H., et al., Monogenic diabetes: a gateway to precision medicine in diabetes. J Clin Invest, 2021. 131(3).

5. Quilichini, E., et al., Insights into the etiology and physiopathology of MODY5/HNF1B pancreatic phenotype with a mouse model of the human disease. J Pathol, 2021. 254(1): p. 31–45.

6. Gardner, D.S. and E.S. Tai, Clinical features and treatment of maturity onset diabetes of the young (MODY). Diabetes Metab Syndr Obes, 2012. 5: p. 101–8.

7. Clissold, R.L., et al., HNF1B-associated renal and extra-renal disease-an expanding clinical spectrum. Nat Rev Nephrol, 2015. 11(2): p. 102–12.

8. Faguer, S., et al., Diagnosis, management, and prognosis of HNF1B nephropathy in adulthood. Kidney International, 2011. 80(7): p. 768–776.

9. Bellanné-Chantelot, C., et al., Clinical spectrum associated with hepatocyte nuclear factor-1beta mutations. Ann Intern Med, 2004. 140(7): p. 510–7.

10. Edghill, E.L., et al., Mutations in hepatocyte nuclear factor-1beta and their related phenotypes. J Med Genet, 2006. 43(1): p. 84–90.

11. Althari, S., et al., Unsupervised Clustering of Missense Variants in HNF1A Using Multidimensional Functional Data Aids Clinical Interpretation. Am J Hum Genet, 2020. 107(4): p. 670–682.

12. Bjørkhaug, L., et al., Hepatocyte Nuclear Factor-1α Gene Mutations and Diabetes in Norway. The Journal of Clinical Endocrinology & Metabolism, 2003. 88(2): p. 920–931.

13. Kind, L., et al., Structural properties of the HNF-1A transactivation domain. Frontiers in Molecular Biosciences, 2023. 10.

14. Artaza, H., et al., Contribution of Rare Large High-Penetrance CNVs to Pediatric and MODY Diabetes in Norway. medRxiv, 2025: p. 2025.02.25.25322584.

15. Faguer, S., et al., The HNF1B score is a simple tool to select patients for HNF1B gene analysis. Kidney Int, 2014. 86(5): p. 1007–15.

16. Nagano, C., et al., Clinical characteristics of HNF1B-related disorders in a Japanese population. Clin Exp Nephrol, 2019. 23(9): p. 1119–1129.

17. Sampathkumar, G., et al., Low genetic confirmation rate in South Indian subjects with a clinical diagnosis of maturity-onset diabetes of the young (MODY) who underwent targeted next-generation sequencing for 13 genes. J Endocrinol Invest, 2022. 45(3): p. 607–615.

18. Domingo-Gallego, A., et al., Clinical utility of genetic testing in early-onset kidney disease: seven genes are the main players. Nephrol Dial Transplant, 2022. 37(4): p. 687–696.

19. Kim, J.H., et al., Etiologic distribution and clinical characteristics of pediatric diabetes in 276 children and adolescents with diabetes at a single academic center. BMC Pediatr, 2021. 21(1): p. 108.

20. Sztromwasser, P., et al., A cross-sectional study of patients referred for HNF1B-MODY genetic testing due to cystic kidneys and diabetes. Pediatr Diabetes, 2020. 21(3): p. 422–430.

21. Monogenic diabetes mellitus in Norway.

22. Gonc, E.N., et al., HNF1B mutation in a Turkish child with renal and exocrine pancreas insufficiency, diabetes and liver disease. Pediatr Diabetes, 2012. 13(2): p. e1–5.

23. Haldorsen, I.S., et al., Lack of pancreatic body and tail in HNF1B mutation carriers. Diabet Med, 2008. 25(7): p. 782–7.

24. Bellanné-Chantelot, C., et al., Large genomic rearrangements in the hepatocyte nuclear factor-1beta (TCF2) gene are the most frequent cause of maturity-onset diabetes of the young type 5. Diabetes, 2005. 54(11): p. 3126–32.

25. Heidet, L., et al., Spectrum of HNF1B mutations in a large cohort of patients who harbor renal diseases. Clin J Am Soc Nephrol, 2010. 5(6): p. 1079–90.

26. Bergmann, C., et al., Mutations in multiple PKD genes may explain early and severe polycystic kidney disease. J Am Soc Nephrol, 2011. 22(11): p. 2047–56.

27. Raaijmakers, A., et al., Criteria for HNF1B analysis in patients with congenital abnormalities of kidney and urinary tract. Nephrol Dial Transplant, 2015. 30(5): p. 835–42.

28. Lindner, T.H., et al., A novel syndrome of diabetes mellitus, renal dysfunction and genital malformation associated with a partial deletion of the pseudo-POU domain of hepatocyte nuclear factor-1beta. Hum Mol Genet, 1999. 8(11): p. 2001–8.

29. Richards, S., et al., Standards and guidelines for the interpretation of sequence variants: a joint consensus recommendation of the American College of Medical Genetics and Genomics and the Association for Molecular Pathology. Genet Med, 2015. 17(5): p. 405–24.

30. Brnich, S.E., et al., Recommendations for application of the functional evidence PS3/BS3 criterion using the ACMG/AMP sequence variant interpretation framework. Genome Med, 2019. 12(1): p. 3.

31. Kołbuc, M., et al., Hyperuricemia Is an Early and Relatively Common Feature in Children with HNF1B Nephropathy but Its Utility as a Predictor of the Disease Is Limited. J Clin Med, 2021. 10(15).

32. Sagen, J.V., et al., Diagnostic screening of MODY2/GCK mutations in the Norwegian MODY Registry. Pediatr Diabetes, 2008. 9(5): p. 442–9.

33. Pavithram, A., et al., In Vitro Functional Analysis Can Aid Precision Diagnostics of HNF1B-MODY. The Journal of Molecular Diagnostics, 2024. 26(6): p. 530–541.

34. The Clinical Genome Resource (ClinGen): Advancing genomic knowledge through global curation. Genet Med, 2025. 27(1): p. 101228.

35. Cheng, J., et al., Accurate proteome-wide missense variant effect prediction with AlphaMissense. Science, 2023. 381(6664): p. eadg7492.

