## Supplementary material for "HNF1B-MODY in the Norwegian MODY Registry and the Norwegian Childhood Diabetes Registry: Clinical insights and prevalence informed by genetic and functional evaluation"

#### Appendix

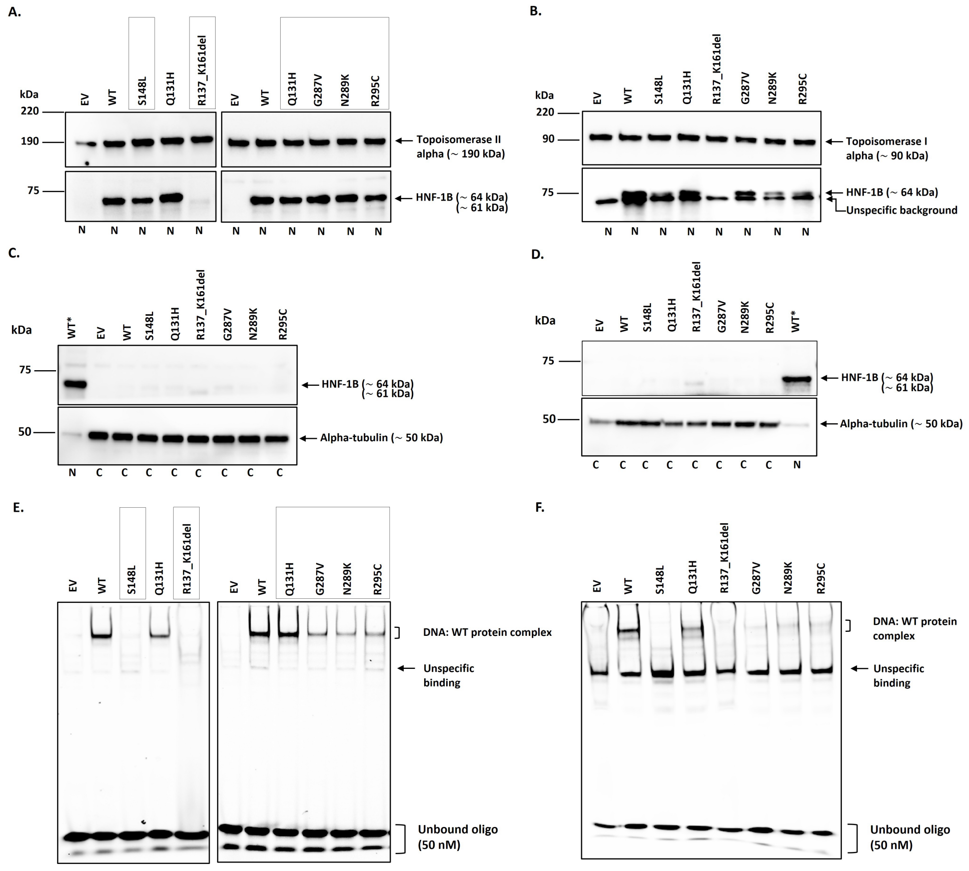


**Supplementary Figure 1.** DNA binding capacity of HNF-1B protein variants. Cells were transfected with EV, WT, or *HNF1B* variant plasmids and lysed 24- or 48-hour post-transfection in HeLa or MIN6 cells. Equal amounts of protein (5 µg) from cytosol and nuclear fraction were analyzed using SDS-PAGE and western blotting. Full-length HNF-1B was detected at ~ 64 kDa, while the deletion variant band is expected to migrate at ~ 61 kDa. Topoisomerase II alpha for HeLa, Topoisomerase I alpha for MIN6 were incorporated as loading controls for nuclear samples and a cytosolic marker, alpha-tubulin is used for cytosol samples. WT* indicates a WT nuclear fraction included in the cytosol panel as an antibody control for HNF-1B detection. Representative blots for nuclear (**A, B**) and cytosolic (**C, D**) fractions are shown for HeLa and MIN6 cells, respectively. Nuclear and cytosol fractions were assessed in three biological replicates (n = 3). Equal amounts of nuclear lysates from HeLa (5 µg) and MIN6 (10 µg) cells were incubated with an oligonucleotide containing the consensus RA-binding site together with EMSA components, and the samples were resolved on retardation gels. Representative EMSA blots from HeLa and MIN6 cells are shown in (**E**) and (**F**). Boxed variant panels in the HeLa nuclear fractions (A) and HeLa EMSA blots represent the signals selected for densitometric analysis and were used for the final quantitative EMSA graph. EMSA runs were performed in three biological replicates in both cell lines (n = 3). EV = Empty vector. WT = wild type.

**Supplementary Table 1. Detailed ACMG-AMP evidence applied for *HNF1B* variants**

| Amino acid Change | Nucleotide Change | Combined ACMG-AMP Evidence | ACMG-AMP Class |
|---|---|---|---|
| p.V2L | c.4G>C | PM1_Supp, PM2_Supp, BS3_Supp | VUS |
| p.S7Rfs*7 | c.18del | PVS1, PP1, PM2_Supp, PP4 | P |
| p.G20R | c.58G>A | PM1_Supp, PP3, PM2_Supp, PP1_Mod, PS4_Mod, PS3_Supp | LP |
| p.L48Rfs*77 | c.143del | PVS1, PM2_Supp, PS2/PM6_Mod, PP4 | P |
| p.P60R | c.179C>G | BS1, BS3_Supp | VUS -> LB |
| p.Q131H | c.393A>T | PM1_Supp, PM2_Supp, PP3 | VUS |
| p.R137_K161del | c.410_484del | PM2_Supp, PM4_Mod, PP1_Mod, PP4, PM1_Mod, PS3_Supp | LP |
| p.Q182* | c.544C>T | PVS1, PS4_Mod, PM2_Supp, PP4 | P |
| p.Q243* | c.727C>T | PVS1, PM2_Supp, PP4 | P |
| p.R276Qfs*51 | c.827del | PVS1, PP4, PM2_Supp | P |
| p.G287V | c.860G>T | PP1_Strong, PM1_Supp, PM2_Supp, PP3, PP4, PS3_Supp | LP |
| p.N289K | c.867C>G | PS4_Mod, PP1, PM1_Supp, PM2_Supp, PM5_Supp, PP4, PS3_Supp | LP |
| p.R295C | c.883C>T | PM1, PM2_Supp, PP3, PS4_Mod, PP1_Strong, PS2/PM6_Supp, PP4, PS3_Supp | P |
| p.N327K | c.981C>G | BS1, BS3_Supp | VUS -> LB |
| p.P343S | c.1027C>T | BP4, BS3_Supp | VUS -> LB |
| p.S362F | c.1085C>T | BS3_Supp | VUS |
| **Controls** | | | |
| p.T186= | c.558A>G | BP7, BS3_Supp | VUS -> LB |
| p.V413= | c.1239C>T | BS1, BP7, BS3_Supp | LB |
| p.S148L | c.443C>T | PS4, PS2/PM6_VS, PM1, PM2_Supp, PP4, PP3, PS3_Supp | P |
| p.R177* | c.529C>T | PVS1, PM2_Supp, PP4 | P |

[§]As per ACMG-AMP, PVS1 (predicted NMD) is applied and not PS3_supp to avoid overweighting the same LoF mechanism.

[†]The p.S148L variant observed in individuals of the NMR cohort and was incorporated as a well-established pathogenic reference variant in the functional assays

BS1 = benign strong. BS3_Supporting = benign supporting. BP2, BP4, BP5, BP7 = benign supporting. PVS1 = pathogenic very strong. VS = Very strong. PS2 = pathogenic strong. PS2/PM6_Supporting = pathogenic supporting. PS2/PM6_Moderate = pathogenic moderate. PS3_Supporting = pathogenic supporting. PS4 = pathogenic strong. PS4_Moderate = pathogenic moderate. PM1, PM2, PM4 = pathogenic moderate. PM1_Supporting, PM2_Supporting = pathogenic supporting. PM4_Moderate = pathogenic moderate. PP1, PP3 = pathogenic supporting. PP1_Moderate = pathogenic moderate. PP1_Strong = pathogenic strong. LP = likely Pathogenic. P = pathogenic. VUS = variants of uncertain significance. LB = Likely benign. B = benign

**Supplementary Table 2. Forward and reverse primer sequences for 17 *HNF1B* variants and controls**

| Variants | Forward Primer Sequence (5' - 3') | Reverse Primer Sequence (5' - 3') |
|---|---|---|
| p.V2L (c.4G>C) | CGATAAGGTACCCATG**C**TGTCCAAGCCACGTC | GACGTGAGCTTGGACA**G**CATGGGTACCTTATCG |
| p.S7R*7 (c.18del) | AGACGCTGCCCCGTCCCCTGGCAG | CTGCCAGGGGACGGGGCAGCGTCT |
| p.G20R (c.58G>A) | CCTGCTGAGCTCC**A**GGGTCACCAAGGA | TCCTTGGTGACCC**T**GGAGCTCAGCAGG |
| p.L48R*77 (c.143del) | GGGTTACATGCAGCATCACAACATCCCCCAG | CTGGGGGATGTTGTGATGCTGCATGTAACCC |
| p.P60R (c.179C>G) | GCCCGACACCAAGC**G**GGTCTTCCATACTC | GAGTATGGAAGACC**C**GCTTGGTGTCGGGC |
| p.Q131H (c.393A>T) | GGGTTACATGCAGCA**T**CACAACATCCCCCAG | CTGGGGGATGTTGTG**A**TGCTGCATGTAACCC |
| p.R137_K161del<br>(c.410_484del) | AGCAACCAAACATCCCCAGACCCAGAAGCGT | AGGCTTCTGGGTCTGGGGATGTTGTTGCT |
| p.S148L (c.443C>T) | CACCGGCCTGAACCAGT**T**GCACCTCTC | GAGAGGTGC**A**ACTGGTTCAGGCCGGTG |
| p.R177* (c.544C>T) | CGACAATTCAACCAGACGGTC-<br>CAGAGTTCTGGAAA | TTTCCAGAACTCTGGACCGTCTGGTTGAATTGTCG |
| p.Q182* (c.544C>T) | GGGGCCCGCGTCC**T**AGCAAATCTTGTAC | GTACAAGATTTGCT**A**GGACGCGGGCCCC |
| p.T186= (c.558A>G) | CGACAATTCAACCAGAC**G**GT<br>CAGAGTTCTGGAAA | TTTCCAGAACTCTGGAC**C**GTCTGGTTGAATTGTCG |
| p.Q243* (c.727C>T) | CCCACGGCCTGGTCTCCAACTTGGTC | GACCAAGTTGGAGACCAGGCCGTGGG |
| p.R276Q*51 (c.827del) | GGCCTGGGCTCCAAGTTGGTCACTGAG | CTCAGTGACCAACTTGGAGCCCAGGCC |
| p.G287V (c.860G>T) | CCCACGGCCTGG**T**CTCCAACTTGGTC | GACCAAGTTGGAG**A**CCAGGCCGTGGG |
| p.N289K (c.867C>G) | GGCCTGGGCTCCAA**G**TTGGTCACTGAG | CTCAGTGACCAA**C**TTGGAGCCCAGGCC |
| p.R295C (c.883C>T) | ACTTGGTCACTGAGGTC**T**GTGTCTACAACTGGTTT | AAACCAGTTGTAGACAC**A**GACCTCAGTGACCA<br>AGT |
| p.N327K (c.981C>G) | GACTCACAGCCTGAA**G**CCTCTGCTCTCCC | GGGAGAGCAGAGG**C**TTCAGGCTGTGAGTC |
| p.P343S (c.1027C>T) | CAGCCCAGCTCCTCT**T**CTCCAAACAAGCTGT | ACAGCTTGTTTGGAG**A**AGAGGAGCTGGGCTG |
| p.S362F (c.1085C>T) | GCAGGGAAACAATGAGATCACTT**T**CTCCT-<br>CAACAATCA | TGATTGTTGAGGAG**A**AAGTGATCTCATT-<br>GTTTCCCTGC |
| p.V413= (c.1239C>T) | GAGGAGGTTTGCCCCCAGT**T**AGCACCTTGA | TCAAGGTGCT**A**ACTGGGGGCAAACCTCCTC |

**Supplementary Table 3. Overview of ACMG-AMP criteria used in the study**

| ACMG-weights | HNF1B-specific Weights | Specifications | Thresholds |
|---|---|---|---|
| | | Benign criteria | |
| BS1 | BS1 | | Grpmax FAF ≥ 1/30,000 (0.000033 or 0.0033%) |
| BS3 | BS3_Supporting | Transactivation assay | ≥ 85% |
| BP4 | BP4 | AlphaMissense scores | ≤ 0.34 |
| BP7 | BP7 | Variant is synonymous with no splice impact, and the nucleotide is weakly/moderately conserved | |
| | | Pathogenic criteria | |
| PVS1 | PVS1 | Variants sensitive to NMD with expected LoF | |
| PS4 | PS4 | Variant met PM2_Supp and observed in multiple cases | 7 or more |
| | PS4_Moderate | | 4-6 |
| PS3 | PS3_Supporting | Impaired transactivation and DNA-binding activity | ≤ 50% |
| PM1 | PM1 | Residues in direct contact with DNA, and applicable for in-frame deletion variants within the DBD | Q136, R137, S142, N146, Q147, S148, H149, N155, K161, K164, R232, R235, K237, Q243, N255, R261, N298, N302, R304, K305 |
| | PM1_Supporting | Residues in critical domains with no direct contact with DNA | DD: 1-30, POU-specific: 101-180, POU-homeo: 229-3[illegible] |
| PM2 | PM2_Supporting | GnomAD v4.1.0 | Grpmax FAF ≤ 1/333,000 (0.000003 or 0.0003%) |
| PM4 | PM4_Moderate | Appliable for in-frame deletion variants | (4-6 aa) |
| PS2 | PS2/PM6_Supporting | Assumed *de novo* and phenotype not specific | |
| | PS2/PM6_Moderate | Assumed *de novo* and phenotype specific | Faguer score ≥ 8 |
| | PS2/PM6_Very strong | Confirmed *de novo* and phenotype specific in two probands | Faguer score ≥ 8 |
| PP1 | PP1 | | 1 family (3 meiosis) and > 1 family (2 meiosis) |
| | PP1_Moderate | Phenotype cosegregates with disease | 1 family (4 meiosis) and > 1 family (3 meiosis) |
| | PP1_Strong | | 1 family (5 meiosis) and > 1 family (4 meiosis) |
| PP3 | PP3 | AlphaMissense scores | ≥ 0.564 |
| PP4* | PP4 | Phenotype is consistent with *HNF1B*-disease | Faguer score ≥ 8 |

*According to the Faguer guidelines, MODY is characterized by a young onset of diabetes (< 35 years), a non-obese presentation, and no insulin dependence[1]. However, since most *HNF1B* patients are insulin-dependent, we assigned 4 points if diabetes was diagnosed before the age of 35 with absent autoantibodies. Other criteria remained unchanged unless stated.

§Residues used for application of the PM1 criterion were based on a previously published study[2]

All variants were classified based on MDEP HNF-1A specific ACMG-AMP variant interpretation guidelines (clinicalgenome.org/affiliation/50016), adapted for the *HNF1B* gene for BS3, BP4, PS3, PM1, PS2, PP3, and PP4. BS1 = benign strong. BS3_Supporting = benign supporting. BP2, BP4, BP5, BP7 = benign supporting. PVS1 = pathogenic very strong. PS2 = pathogenic strong. PS2/PM6_Supporting = pathogenic supporting. PS2/PM6_Moderate = pathogenic moderate. PS3_Supporting = pathogenic supporting. PS4 = pathogenic strong. PS4_Moderate = pathogenic moderate. PM1, PM2, PM4 = pathogenic moderate. PM1_Supporting, PM2_Supporting = pathogenic supporting. PM4_Moderate = pathogenic moderate. PP1, PP3 = pathogenic supporting. PP1_Moderate = pathogenic moderate. PP1_Strong = pathogenic strong.

### References

1. Faguer, S., et al., *The HNF1B score is a simple tool to select patients for HNF1B gene analysis.* Kidney Int, 2014. **86**(5): p. 1007-15.
2. Lu, P., G.B. Rha, and Y.I. Chi, *Structural basis of disease-causing mutations in hepatocyte nuclear factor 1beta.* Biochemistry, 2007. **46**(43): p. 12071-80.
